# Cardiac Magnetic Resonance Findings of COVID-19 Vaccine Associated Myocarditis at Intermediate Follow Up: a Comparison to Classic Myocarditis and MIS-C Related Myocarditis

**DOI:** 10.1101/2022.09.01.22279517

**Authors:** Matthew L. Dove, Timothy C. Slesnick, Matthew E. Oster, Sassan Hashemi, Trisha Patel, Hunter C. Wilson

**Affiliations:** Department of Pediatrics, Division of Cardiology, Emory University School of Medicine, Children’s Healthcare of Atlanta, Atlanta, GA

**Keywords:** CMR, C-VAM, SARS-CoV-2, COVID vaccine, BNT162 vaccine, mRNA vaccine

## Abstract

**Objective:** To report intermediate cardiac magnetic resonance (CMR) findings of COVID-19 vaccine associated myocarditis (C-VAM) and compare to classic myocarditis (CM) and multisystem inflammatory syndrome in children (MIS-C).

**Study Design:** Retrospective cohort study including children diagnosed with C-VAM from 5/2021 through 12/2021 with early and intermediate CMR. Patients with CM and MIS-C with intermediate CMR were included for comparison.

**Results:** There were 8 patients with C-VAM, 20 with CM, and 61 with MIS-C. Among those with C-VAM, CMR performed at median 3 days (IQR 3, 7) revealed 2/8 patients with left ventricular ejection fraction (LVEF)<55%, 7/7 patients with late gadolinium enhancement (LGE), and 5/8 patients with elevated native T1 values. Borderline T2 values suggestive of myocardial edema were present in 6/8. Follow-up CMRs performed at median 107 days (IQR 97, 177) showed normal ventricular systolic function, T1, and T2 values; 3/7 patients had LGE. At intermediate follow up the C-VAM group had a lower percentage of LVEF<55% compared to CM and MIS-C (0.0 vs 30.0 vs 6.6%, respectively, p=0.018) and an intermediate degree of LGE (42.9 vs 75.0 vs 3.3%, respectively, p<0.001). Pairwise comparisons showed fewer myocardial segments with LGE in the C-VAM group versus CM (4/119 vs 42/340, p=0.004) and more segments with LGE than MIS-C (4/119 vs 2/1020, p=0.0014).

**Conclusion:** Patients with C-VAM had no evidence of active inflammation or ventricular dysfunction on intermediate CMR although a minority had persistent LGE. Intermediate findings in C-VAM may be favorable compared to CM though LGE is more common compared to MIS-C.

## Introduction

Myocarditis has been identified as a rare but serious adverse event following mRNA COVID-19 vaccination, predominantly affecting adolescent males. These cases are characterized by chest pain, troponin elevation, and ST segment or T wave changes on electrocardiogram within several days of receiving the vaccine. Cardiac magnetic resonance (CMR) imaging obtained during the acute illness frequently reveals late gadolinium enhancement (LGE) and myocardial edema(1-6). Classic viral myocarditis and multisystem inflammatory syndrome in children (MIS-C) are two other illnesses that demonstrate CMR evidence of myocardial inflammation. Many children with classic myocarditis have ventricular systolic dysfunction and myocardial fibrosis that persist beyond the acute phase(7-9) whereas these findings largely resolve in children with MIS-C(10, 11). While the short-term outcomes of COVID-19 vaccine associated myocarditis (C-VAM) have been shown to be less severe than those with classic viral myocarditis or MIS-C(12), the intermediate-term effects are unknown. We aimed 1) to describe the CMR findings of COVID-19 vaccine associated myocarditis patients at intermediate follow up and 2) to compare these findings to those of children with classic myocarditis and MIS-C.

## Methods

### Study Design and Patient Selection

We performed a retrospective cohort study of patients aged <21 years diagnosed with C-VAM at our institution between May 2021 and December 2021. Patients who had an CMR at presentation and underwent follow-up CMR were included. Patients who received a follow-up CMR 2-12 months after diagnosis of viral or idiopathic myocarditis from January 2015 to December 2021 or 2-12 months after diagnosis of MIS-C from April 2020 to June 2021 were included for the comparison groups. CMR findings in a portion of the latter group were previously reported; the group was expanded to include additional patients to generate the comparator group used for the current study(10). We required negative COVID-19 PCR and IgG anti-nucleocapsid antibody testing for patients who experienced classic myocarditis in 2020 and later to qualify for inclusion in the classic myocarditis group. Demographic, clinical, echocardiographic data, and CMR data were collected from time of hospitalization through follow-up CMR for the C-VAM group. Demographic and CMR data were collected for the classic myocarditis and MIS-C groups. Comparison of functional data, volumetric data, parametric mapping, and LGE were made among the C-VAM, classic myocarditis, and MIS-C groups. The institutional review board at our institution approved this study.

### CMR Acquisition and Analysis

All CMR examinations were performed on a 1.5 Tesla magnet (Avanto Fit, Siemens Healthcare, Erlangen, Germany). Most studies were performed awake with breath holding, while a small subset of patients required general anesthesia due to younger age; in these patients, free breathing techniques with multiple signal averages were used. All studies were performed with administration of 0.2 mmol/kg intravenous gadolinium contrast (gadobutrol, Gadavist, Bayer Healthcare Pharmaceuticals, Wayne, NJ). Pre-contrast steady-state free precession cine imaging was performed in the 2-chamber, 3-chamber, 4-chamber, and short axis geometries, and left and right ventricular volumes, ejection fraction, and left ventricular mass were recorded. Pre- and post-contrast T1 mapping as well as pre-contrast T2 mapping were performed in the short axis geometry at the basilar, mid, and apical levels. Standard modified look-locker inversion recovery (MOLLI) sequences were used, with a 5(3)3 acquisition strategy for pre-contrast T1 and a 4(1)3(1)2 sequence for the post-contrast T1 imaging. Parametric mapping has been favored over traditional T1- and T2-weighted black blood imaging at our center for tissue characterization for the past several years but was less frequently performed in earlier studies. LGE imaging was performed in both the short axis and 4-chamber geometries with single shot phase sensitive inversion recovery imaging and a look-locker sequence used to set the inversion time.

Post-processing readings were performed by a single observer with eight years of post-processing experience (SH) who was blinded to the clinical data. Ventricular volumes, mass, and function were calculated from short axis cines. T1 pre, T1 post, and T2 maps had regions of interest (ROIs) drawn on both the septum and free wall at the apex, mid, and basilar levels, in addition to an ROI in the blood pool for extracellular volume (ECV) calculations. Only mid and basilar values were included in this study given increased imaging heterogeneity in apical segments, especially in younger patients. In addition to these ROIs, if there was an area of focal abnormality on any of the T1 or T2 maps, that area was also contoured for comparison to the rest of the myocardium. The modified Lake Louise criteria were used to assess for myocarditis(13). ECV was calculated using the patient’s last recorded hematocrit. If no hematocrit was available within one month of the CMR study, an estimated hematocrit of 40% was used. In our lab, upper limits of normal for native T1 is defined at 1050ms, T2 at 60ms, and ECV at 30%. LGE was assessed and described as either present or absent in each of the seventeen segments in the standard American Heart Association model for each patient(14). LGE imaging was also qualitatively reviewed by a single user (HW) for each C-VAM and classic myocarditis patient. When LGE location and characterization were not discretely specified in CMR reports, images were reviewed and locations and characterization of LGE patterns were assigned.

### Statistical Analysis

Statistical analysis was completed using SPSS Statistics version 28.0.0.0. Descriptive data are presented as medians with interquartile ranges for continuous data and counts and percentages for categorical data. Continuous data were compared using the Kruskal-Wallis test. Categorical variables were compared using Fisher’s exact test. A *p*-value of less than 0.05 was considered significant. Post-hoc comparison of patient groups was performed by including only patients ≥ 12 years of age given that the COVID-19 vaccine was not approved for children younger than 12 years of age for the majority of the study period. Post-hoc testing was also completed for variables of interest by comparing patients with C-VAM to classic myocarditis and to MIS-C groups individually using Fisher’s exact test.

## Results

### C-VAM outcomes

The C-VAM group included 8 patients. They were all male, and had a median age of 16 years (IQR 15, 17) at time of diagnosis. All patients received the Pfizer-BioNTech COVID-19 vaccine (the only COVID-19 vaccine authorized for those aged 12-17 years at the time of initial enrollment for this study), with 2 children developing myocarditis after the first dose and 6 after the second dose. Median time from vaccination to diagnosis was 3 days (IQR 3, 7). All children reported chest pain and had an elevated troponin (upper limit of normal <0.045 ng/mL) with median peak troponin of 6.15 ng/mL (IQR 4.37, 9.89). By echocardiogram, the average left ventricular ejection fraction (LVEF) at time of presentation was 63.3 ± 11.5%, with 2/8 having LVEF <55%. Both patients had normalization of LVEF within 13 days. Two patients were admitted to the intensive care unit, one patient received inotropic infusions, and all eight were treated with non-steroidal anti-inflammatory drugs. Median length of hospitalization was 2 days (IQR 1, 3).

Among those with C-VAM, initial CMR revealed 2/8 patients with LVEF <55%, 7/7 patients who received contrast with LGE indicating myocardial fibrosis, 5/8 patients with elevated native T1 values, and 4/7 with elevated ECV (Table 1). No patient had an elevated T2 value per the institutional cutoff of 60ms; accordingly, there were no patients who met strict criteria for myocarditis using Lake Louise criteria. However, T2 mapping revealed a heterogenous myocardium and ROIs with borderline elevated T2 values relative to surrounding myocardium in 6/8 children, suggestive of myocardial edema. LGE predominantly involved the inferior and inferolateral segments of the heart (Figure 1). 3/7 children had a subepicardial and mid-myocardial pattern of LGE while 4/7 had a subepicardial pattern only. Intermediate follow-up CMR was performed a median of 107 days (97, 177) after diagnosis. 3/7 patients had persistent subepicardial LGE, and all patients had normal ventricular systolic function, T1, ECV, and T2 values (Figure 2). There was one patient whose parents declined contrast on both initial and follow up CMR. This patient’s initial CMR showed elevated basal and mid T1 relaxation times in a subepicardial distribution in the inferolateral segments and borderline T2 elevation relative to surrounding myocardium which normalized on follow-up CMR.

**Table 1.** Early and intermediate CMR findings amongst children with C-VAM

|  | Baseline CMR (n=8) | Follow-up CMR (n=8) | P-value |
| --- | --- | --- | --- |
| Time since diagnosis (d) | 3 (3, 7) | 107 (97, 177) | **<0.001** |
| LVEF (%) | 56.5 (54.8, 59.8) | 57.0 (55.8, 60.5) | 0.597 |
| LVEF <55% | 2/8 (25.0) | 0/8 (0.0) | 0.467 |
| RVEF (%) | 52.5 (49.0, 51.0) | 52.5 (51.0, 53.5) | 0.711 |
| Native T1 base (ms) | 1038.4 (1002.4, 1053.1) | 990.3 (985.1, 1017.8) | **0.046** |
| Native T1 mid (ms) | 1012.7 (980.7, 1031.5) | 984.5 (976.0, 1004.5) | 0.115 |
| Native T1 ROI (ms) | 1143.3 (1040.0, 1160.2) n=5 | - | N/A |
| Native T1 >1050ms<sup>α</sup> | 5/8 (62.5) | 0/8 (0.0) | **0.026** |
| T2 base (ms) | 50.2 (46.0, 51.5) | 47.3 (46.3, 48.0) | 0.269 |
| T2 mid (ms) | 49.2 (47.7, 50.3) | 47.6 (47.0, 48.0) | 0.074 |
| T2 ROI (ms) | 56.0 (54.8, 57.9) n=6 | - | N/A |
| T2 >60ms<sup>α</sup> | 0/8 (0.0) | 0/8 (0.0) | N/A |
| ECV Base (%) | 24.2 (22.9, 27.5) | 25.2 (24.3, 25.8) | 0.873 |
| ECV Mid (%) | 23.6 (21.0, 27.3) | 25.4 (22.1, 26.5) | 0.749 |
| ECV ROI | 31.4 (26.3, 36.9) n=4 | - | N/A |
| ECV >30%<sup>α</sup> | 4/7 (57.1) n=7 | 0/6 (0.0) n=6 | 0.070 |
| LGE Present | 7/7 (100.0) n=7 | 3/7 (42.9) n=7 | 0.0699 |
| Total segments with LGE | 13/119 (10.9) n=7 | 4/119 (3.4) n=7 | **0.0412** |

Data are presented as Median (Interquartile Range) or N (%).
<sup>α</sup> Each count represents patient with value elevated above specified limit in either basal LV, mid LV, or region of interest

**Figure 1.**
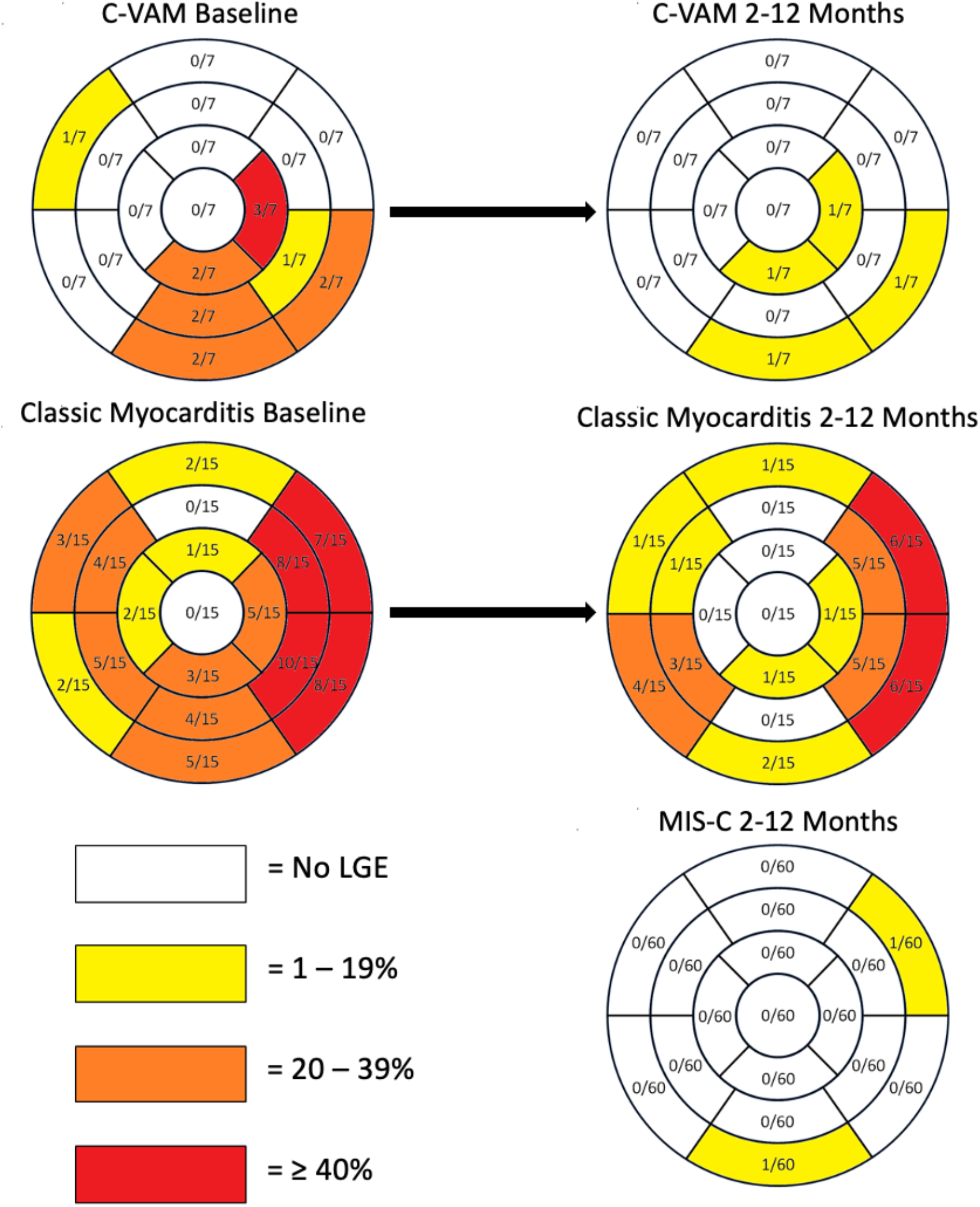
LGE distribution and burden among myocarditis groups Frequency of left ventricular wall segments with LGE among patients with C-VAM, classic myocarditis, and MIS-C. Segments are recorded in accordance with the 17-segment AHA heart model. The classic myocarditis 2-12 month graphic only includes the 15 classic myocarditis patients who had early CMR.

**Figure 2.**
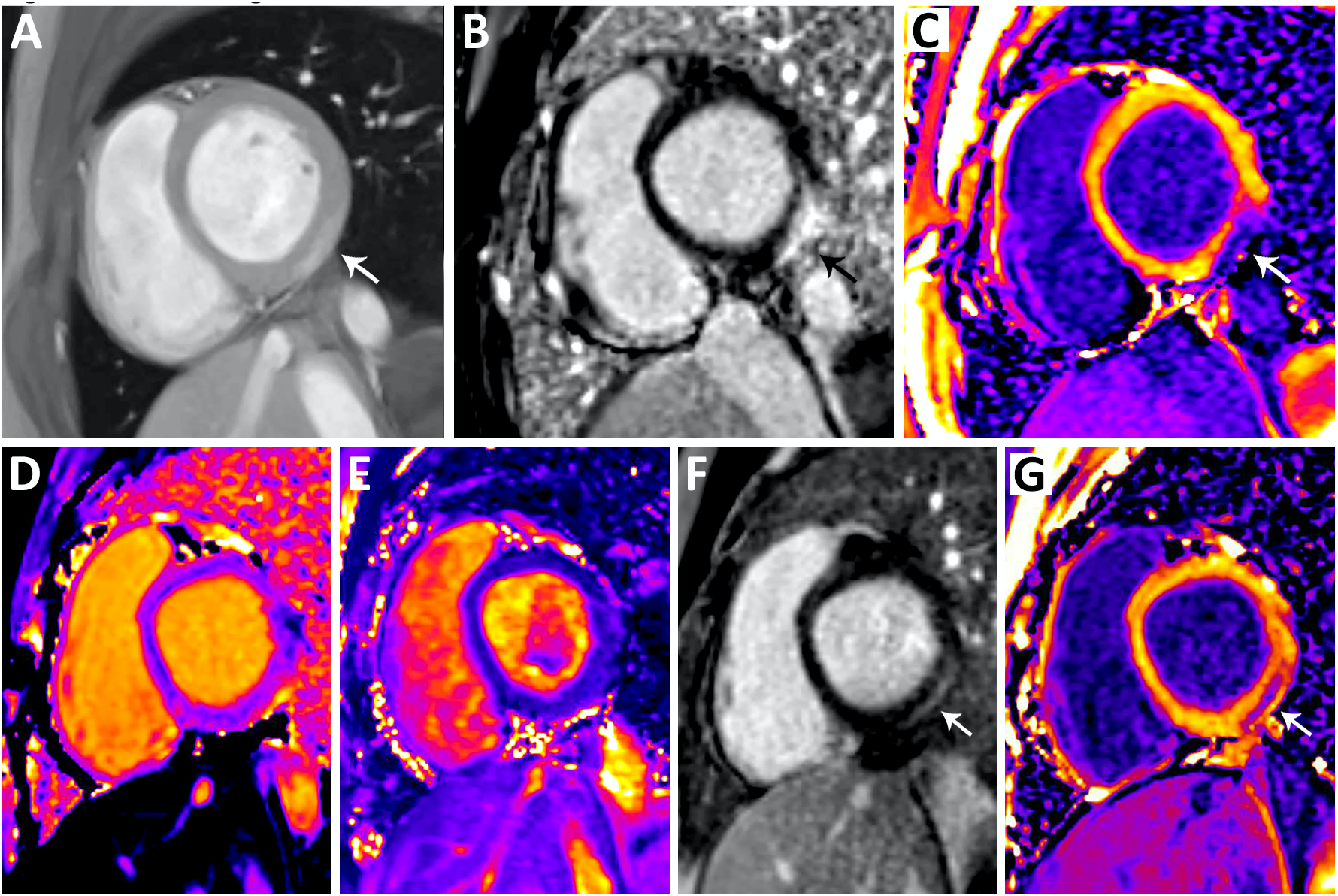
CMR Images from C-VAM Patient Top row: Baseline CMR performed 1 day after diagnosis of C-VAM with short axis images including (A) early post-contrast T1-weighted spoiled gradient compressed SENSE cine still frame showing regional signal hyperintensity (B) late gadolinium enhancement and (C) corresponding regional post-contrast shortening of T1 relaxation times in the basilar inferolateral segment as depicted by the arrow. Bottom row: Follow-up CMR performed at 89 days after diagnosis with short axis images including (D) normal native T1 mapping relaxation times (E) normal T2 mapping relaxation times (F) persistent late gadolinium enhancement and (G) post-contrast shortening of T1 relaxation times in the basilar inferolateral segment as depicted by the arrow.

### Comparison of C-VAM outcomes to classic myocarditis and MIS-C myocarditis outcomes

20 children with classic myocarditis and 61 children with MIS-C myocarditis were included for the comparison groups (Table 2). Patients with C-VAM and classic myocarditis were older and more frequently male than those with MIS-C. The majority of patients in the C-VAM group were non-Hispanic White, whereas the majority of patients in the classic myocarditis and MIS-C group were non-Hispanic Black. Mean native T1 values were significantly different between groups, with the longest relaxation times in the MIS-C group; T2 values and ECV values were not significantly different among groups. The classic myocarditis group had a higher percentage of patients with LVEF <55% than the MIS-C and C-VAM groups (30.0% vs 6.6% vs 0.0%, respectively, p=0.018) and RVEF <47% (15.0% vs 0.0% vs 0.0%, respectively, p=0.015). Amongst all groups, no patient had evidence of myocardial edema or met the modified Lake Louise criteria for myocarditis. The frequencies of LGE among groups were significantly different, with the highest frequency of LGE noted in the classic myocarditis group compared to the MIS-C and C-VAM groups (75.0% vs 3.3% vs 42.9%, respectively, p<0.001). Pairwise comparisons of C-VAM patients with classic myocarditis patients showed no significant differences in percentages of patients with abnormal LVEF, RVEF, or LGE, although the percentage of myocardial segments with LGE among all patients was significantly lower in the C-VAM group (3.4 vs 12.3%, p=0.004). When similarly compared with MIS-C patients, the percentages of C-VAM patients with abnormal ventricular function were not significantly different, whereas the percentage of patients with LGE was higher (42.9 vs 3.3%, p=0.0066) as was the total percentage of myocardial segments with LGE among all patients (3.4 vs 0.2%, p=0.0014) (Supplement 1). The LGE pattern was subepicardial in the C-VAM group compared to a more diverse pattern in the classic myocarditis group (Supplement 2). Among the classic myocarditis group, there were 15 patients who had early CMR performed following diagnosis. Both C-VAM and classic myocarditis patients showed reduction in number of segments with LGE between early and intermediate CMR. Comparison of intermediate CMR among groups showed that LGE was similarly distributed primarily in inferolateral and inferior segments for C-VAM and more widely among the classic myocarditis group although most frequently within the anterolateral and inferolateral segments (Figure 1). LGE among the 15 classic myocarditis patients with early CMR and the full complement of 20 classic myocarditis patients with intermediate CMR is shown in Supplement 3.

**Table 2.** Comparison of demographic and CMR Findings at Intermediate Follow Up

|  | Vaccine Myocarditis<br>(N=8) | Classic Myocarditis<br>(N=20) | MIS-C Myocarditis<br>(N=61) | P-value |
| --- | --- | --- | --- | --- |
| Age (years) | 16.3 (15.7, 17.0) | 16.7 (14.9, 17.5) | 11.9 (8.5, 15.2) | **<0.001** |
| Sex |  |  |  | **0.030** |
| Male | 8/8 (100.0) | 18/20 (90.0) | 41/61 (67.2) |  |
| Female | 0/8 (0.0) | 2/20 (10.0) | 20/61 (32.8) |  |
| Race/Ethnicity |  |  |  | **<0.001** |
| NH White | 6/8 (75.0) | 8/20 (40.0) | 7/61 (11.5) |  |
| NH Black | 1/8 (12.5) | 10/20 (50.0) | 43/61 (70.5) |  |
| Hispanic | 0/8 (0.0) | 2/20 (10.0) | 7/61 (11.5) |  |
| Other | 1/8 (12.5) | 0/20 (0.0) | 4/61 (6.6) |  |
| BSA (m<sup>2</sup>) | 1.86 (1.76, 2.24) | 1.94 (1.71, 2.09) | 1.60 (1.15, 2.09) | **0.018** |
| Time between diagnosis and CMR (days) | 107 (97, 177) | 111 (101, 195) | 105 (93, 145) | 0.415 |
| Anesthesia during case | 0/8 (0.0) | 2/20 (10.0) | 15/61 (24.6) | 0.166 |
| LVEDV/BSA (mL/m<sup>2</sup>) | 73.5 (70.1, 78.1) | 75.3 (66.8, 82.5) | 71.0 (61.0, 77.0) | 0.143 |
| LVESV/BSA (mL/m<sup>2</sup>) | 32.1 (28.0, 34.1) | 33.0 (26.0, 35.3) | 24.0 (24.0, 32.0) | **0.043** |
| LVEF (%) | 57.0 (55.8, 60.5) | 58.0 (54.0, 61.3) | 60.0 (58.0, 63.0) | 0.168 |
| LVEF <55% | 0/8 (0.0) | 6/20 (30.0) | 4/61 (6.6) | **0.018** |
| RVEDV/BSA (mL/m<sup>2</sup>) | 82.0 (78.1, 85.2) | 83.5 (70.4, 92.5) | 75.0 (68.4, 84.4) | 0.228 |
| RVESV/BSA (mL/m<sup>2</sup>) | 37.5 (36.8, 41.7) | 38.7 (29.9, 44.2) | 33.0 (29.1, 37.5) | 0.101 |
| RVEF (%) | 52.5 (51.0, 53.5) | 53.0 (49.0, 56.5) | 55.0 (52.0, 58.0) | 0.130 |
| RVEF <47% | 0/8 (0.0) | 3/20 (15.0) | 0/61 (0.0) | **0.015** |
| Native T1 base (ms) | 990.3 (985.1, 1017.8) | 1002.0 (994.1, 1017.9)<br>n=13 | 1019.0 (1001.0, 1033.5) n=58 | **0.033** |
| Native T1 mid (ms) | 984.5 (976.0, 1004.5) | 997.5 (980.2, 1010.9)<br>n=11 | 1009.7 (992.0, 1032.0)<br>n=50 | 0.071 |
| Native T1 >1050ms<sup>α</sup> | 0/8 (0.0) | 2/14 (14.3) | 10/61 (16.3) | 0.677 |
| Native T2 base (ms) | 47.3 (46.3, 48.0) | 47.6 (45.4, 48.2) n=11 | 47.8 (46.5, 49.2) n=58 | 0.581 |
| Native T2 mid (ms) | 47.6 (47.0, 48.0) | 47.8 (47.0, 49.3) n=10 | 47.7 (46.3, 49.0) n=51 | 0.712 |
| Native T2 >60ms<sup>α</sup> | 0/8 (0.0) | 0/11 (0.0) | 0/59 (0.0) | N/A |
| ECV Base (%) | 25.2 (24.3, 25.8) n=6 | 24.0 (20.9, 26.0) n=12 | 22.9 (21.8, 25.1) n=55 | 0.400 |
| ECV Mid (%) | 25.4 (22.1, 26.5) n=6 | 24.7 (23.4, 24.9) n=11 | 23.4 (22.2, 24.9) n=46 | 0.519 |
| ECV >30%<sup>α</sup> | 0/6 (0.0) | 2/14 (14.3) | 2/59 (3.4) | 0.264 |
| LGE Present | 3/7 (42.9) | 15/20 (75.0) | 2/60 (3.3) | **<0.001** |
| Total segments with LGE | 4/119 (3.4) | 42/340 (12.4) | 2/1020 (0.2) | **<0.001** |

Data are presented as Median (Interquartile Range) or N (%). Patient group sizes as indicated in top row unless otherwise noted by denominator.
<sup>α</sup> Each count represents patient with value elevated above specified limit in either basal LV, mid LV, or region of interest.

After post hoc comparison excluding patients <12 years of age, mean patient age and size were similar, as were native T1 relaxation times, although the percentage of patients with LVEF <55% and persistent LGE remained significantly different between groups (Supplement 4).

## Discussion

In our study of patients with COVID-19 vaccine associated myocarditis, intermediate follow-up CMR demonstrated resolution of inflammation, myocardial edema, and ventricular dysfunction. While improvement in the burden of LGE was seen, some patients had persistent LGE consistent with myocardial fibrosis. Ventricular systolic dysfunction and myocardial fibrosis were most frequent among patients diagnosed with classic viral myocarditis as compared to patients with C-VAM and MIS-C. This is the first study to our knowledge to compare intermediate CMR findings among children and young adults with C-VAM, classic myocarditis, and MIS-C.

Although there was heterogeneity in T2 maps and corresponding T1 abnormalities among early CMR studies in C-VAM patients, no patients exceeded our institutional cutoff of 60ms used to indicate myocardial edema. Thus, no patients satisfied formal Lake Louise criteria for diagnosis of myocarditis, although temporal relationship to immunization, troponin elevation, and symptoms provide strong support for clinical diagnosis of myocarditis. Our findings are similar to those from Dionne et al who reported CMR evidence of myocardial edema in only 13% of patients with C-VAM undergoing acute CMR(15). Notably, the one patient in whom contrast was not administered and LGE could consequently not be assessed did show elevated basal native T1 values in a subepicardial distribution, supporting the value of parametric mapping as a diagnostic tool for myocardial injury even for patients in whom contrast is contraindicated or declined.

The American Heart Association and American College of Cardiology have recommended a minimum interval of 3-6 months after acute myocarditis prior to resumption of sports, given the link to sudden cardiac death(16, 17). This interval was adequate time to allow for normalization of MRIs in at least half of our cohort, with 3 of the remaining patients showing persistant LGE with otherwise normal ventricular systolic function and no other evidence of active myocardial inflammation. Schauer et al and Fronza et al have also shown resolution of myocardial edema and normalization of ventricular systolic function in the same follow-up period(18, 19). These studies also revealed persistent LGE albeit at a higher frequency than our group: 69% and 62%, respectively. A more recent report from Hadley et al comparing early and intermediate CMR in C-VAM patients showed an even higher percentage of patients with LGE in intermediate CMR than we report (80%), although the degree of LGE was improved in comparison to early CMR (20). In the adult population, both the presence and a higher burden of LGE has been associated with increased risk of adverse cardiac outcomes(21). Clinical significance of isolated persistant LGE in C-VAM is unknown, but clinicians may consider framing decisions on sports clearance and follow up in accordance with recommendations which have been issued for patients with classic myocarditis(16, 17, 22). Although ours and other centers’ data support favorable early and intermediate outcomes, further data are needed to better understand long term clinical sequelae of C-VAM.

In our comparison, the classic myocarditis group had a higher burden of LGE than the C-VAM group at intermediate term follow-up. These findings suggest the possibility of a more favorable prognosis in children with C-VAM as opposed to classic myocarditis and extend earlier findings from our group comparing early clinical outcomes among patients with C-VAM, classic myocarditis, and MIS-C(12). The pattern of LGE at intermediate CMR was exclusively subepicardial in our group, compared to the classic myocarditis group, which showed a wider variety in patterns of enhancement. Findings are similar to those from Patel et al, who noted more frequent mid myocardial LGE in classic myocarditis patients compared to C-VAM patients, along with reduced LVEF in the former(23). Although our baseline analysis revealed a significant difference in native T1 values among groups, with a higher T1 relaxation time in children with MIS-C, this finding is clouded by the younger age of the MIS-C cohort. T1 values have been shown to increase with younger age and higher heart rate(24, 25). When we limited our age ranges to patients older than 12 years old, no significant differences between native T1 values were noted, and no patients older than 12 in the MIS-C group had native T1 values above our cutoff of abnormal. While the possibility of microscopic fibrosis in younger patients cannot be excluded, the otherwise generally normal CMR indices in these patients provide indirect support that higher values in younger patients may attributable to patients’ ages rather than indicative of true myocardial fibrosis(10). Furthermore, the degree of LGE in MIS-C patients was minimal and significantly less than among pateints with C-VAM. Notably, abnormal LVEF and frequency of LGE remained significantly different among groups in post-hoc analysis despite similar ages in each group, suggesting that these differences are not linked to patients’ ages. Limitations of this study include inherent variability associated with parametric mapping indices, with prior data showing differences between vendors and sites that may be significant, prompting development of consensus recommendations(26). Some patients in the classic myocarditis group with an earlier diagnosis did not have as extensive parametric data available for comparison. Our study groups for our primary analysis do not include patients without CMR testing and may consequently reflect patients with a more severe phenotype in each group.

Finally, the smaller number of C-VAM patients limits the power of our statistical analysis, although the size of this cohort is similar to those reported in other single center studies. Future directions include long term monitoring for adverse cardiac events and the association with persistent LGE in the C-VAM population.

## Data Availability

All data produced in the present work are contained in the manuscript.

## Abbreviations

BSA: body surface area
CHOA: Children’s Healthcare of Atlanta
CMR: cardiac magnetic resonance
C-VAM: COVID-19 vaccine associated myocarditis
ECV: extracellular volume
LGE: late gadolinium enhancement
LVEDV: left ventricular end diastolic volume
LVEF: left ventricular ejection fraction
MIS-C: Multisystem Inflammatory Syndrome in Children
ROI: region of interest
RVEDV: right ventricular end diastolic volume
RVEF: right ventricular end diastolic volume

---

**Supplement 1.** Pairwise comparisons of C-VAM patients with classic myocarditis patients and MIS-C patients specifically for variables of interest.

| C-VAM versus classic myocarditis |  |  |  |
| --- | --- | --- | --- |
|  | C-VAM | Classic Myocarditis | P-value |
| LVEF < 55% | 0/8 (0.0) | 6/20 (30.0) | 0.141 |
| RVEF < 47% | 0/8 (0.0) | 3/20 (15.0) | 0.536 |
| LGE | 3/7 (42.9) | 15/20 (75.0) | 0.175 |
| Total Segments with LGE | 4/119 (3.4) | 42/340 (12.4) | 0.004 |
| C-VAM versus MIS-C |  |  |  |
|  | C-VAM | MIS-C | P-value |
| LVEF < 55% | 0/8 (0.0) | 4/61 (6.6) | 1.0 |
| RVEF < 47% | 0/8 (0.0) | 0/61 (0.0) | 1.0 |
| LGE | 3/7 (42.9) | 2/60 (3.3) | **0.0066** |
| Total Segments with LGE | 4/119 (3.4) | 2/1020 (0.2) | 0.0014 |

Data are presented as N (%).

**Supplement 2.** LGE patterns on follow-up CMR by group

| **Pattern** | **Vaccine Myocarditis (n=7)** | **Classic Myocarditis (n=20)** | **MIS-C Myocarditis (n=60)** |
| --- | --- | --- | --- |
| Subepicardial | 3 (42.9%) | 10 (50.0%) | 0 (0.0%) |
| Mid myocardial | 0 (0.0%) | 6 (30.0%) | 1 (1.7%) |
| Subendocardial | 0 (0.0%) | 2 (10.0%) | 1 (1.7%) |
| Transmural | 0 (0.0%) | 1 (5.0%) | 0 (0.0%) |

Data are presented as N (%).

**Supplement 3.**
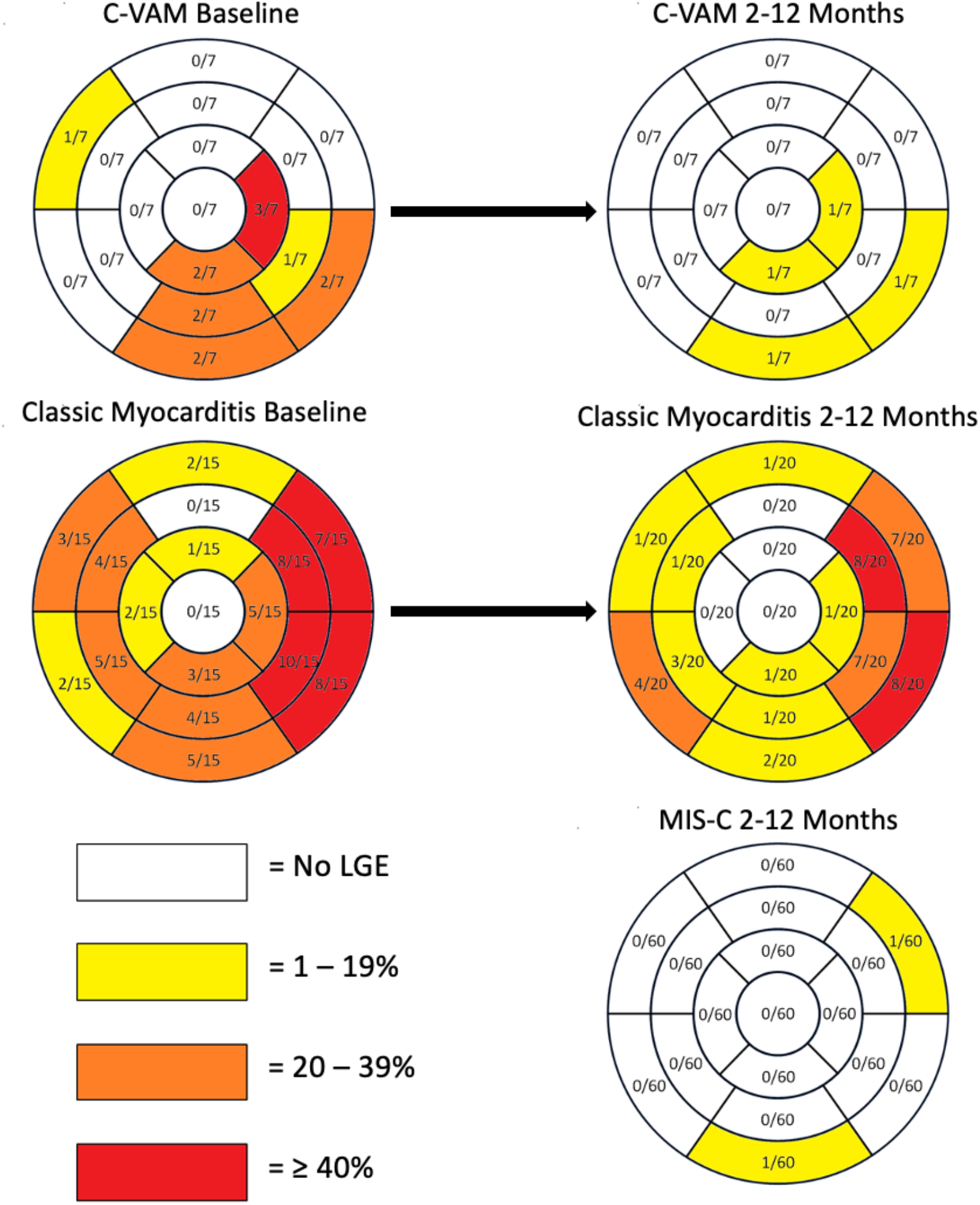
LGE distribution and burden among myocarditis groups with inclusion of the 15 classic myocarditis patients with early CMR and the full complement of 20 patients with intermediate CMR. Frequency of left ventricular wall segments with LGE among patients with C-VAM, classic myocarditis, and MIS-C. Segments are recorded in accordance with the 17-segment AHA heart model. The early classic myocarditis graphic includes 15/20 study patients with early CMR, and the 2-12 month classic myocarditis graphic includes the full complement of 20 patients who had intermediate CMR available.

**Supplement 4.** Sensitivity analysis with exclusion of patients <12 years of age

|  | Vaccine Myocarditis<br>(N=8) | Classic Myocarditis<br>(N=19) | MIS-C Myocarditis<br>(N=29) | P-value |
| --- | --- | --- | --- | --- |
| Age (years) | 16.3 (15.7, 17.0) | 16.4 (14.7, 17.3) | 14.8 (13.5, 16.3) | 0.118 |
| Sex |  |  |  | 0.381 |
| Male | 8/8 (100.0) | 17/19 (89.5) | 23/29 (79.3) |  |
| Female | 0/8 (0) | 2/19 (10.5) | 6/29 (20.7) |  |
| Race/Ethnicity |  |  |  | **0.008** |
| NH White | 6/8 (75.0) | 8/19 (42.1) | 4/29 (13.8) |  |
| NH Black | 1/8 (12.5) | 9/19 (47.4) | 20/29 (69.0) |  |
| Hispanic | 0/8 (0) | 2/19 (10.5) | 3/29 (10.3) |  |
| Other | 1/8 (12.5) | 0/19 (0) | 2/29 (6.9) |  |
| BSA (m<sup>2</sup>) | 1.86 (1.76, 2.24) | 1.95 (1.77, 2.11) | 1.95 (1.80, 2.17) | 0.927 |
| Time between diagnosis and CMR (days) | 107 (97, 177) | 106 (102, 199) | 105 (94, 142) | 0.512 |
| Anesthesia during case | 0/8 (0) | 1/19 (5.3) | 0/29 (0) | 0.482 |
| LVEDV/BSA (mL/m<sup>2</sup>) | 73.5 (70.1, 78.1) | 78.0 (66.8, 82.7) | 76.0 (66.0, 81.8) | 0.936 |
| LVESV/BSA (mL/m<sup>2</sup>) | 32.1 (28.0, 34.1) | 33.0 (27.3, 35.5) | 29.7 (26.0, 32.0) | 0.312 |
| LVEF (%) | 57.0 (55.8, 60.5) | 58.0 (54.0, 61.5) | 60.0 (58.0, 63.0) | 0.181 |
| LVEF <55% | 0/8 (0) | 6/19 (31.6) | 1/29 (3.4) | **0.012** |
| RVEDV/BSA (mL/m<sup>2</sup>) | 82.0 (78.1, 85.2) | 84.0 (69.8, 93.0) | 84.0 (75.0, 92.1) | 0.900 |
| RVESV/BSA (mL/m<sup>2</sup>) | 37.5 (36.8, 41.7) | 39.0 (30.8, 45.4) | 36.0 (33.0, 44.0) | 0.947 |
| RVEF (%) | 52.5 (51.0, 53.5) | 52.0 (49.0, 55.5) | 54.0 (51.0, 58.0) | 0.410 |
| RVEF <47% | 0/8 (0) | 3/19 (15.8) | 0/29 (0) | 0.085 |
| Native T1 base (ms) | 990.3 (985.1, 1017.8) | 1001.5 (991.1, 1011.8)<br>n=12 | 1003.2 (987.9, 1017.5)<br>n=28 | 0.827 |
| Native T1 mid (ms) | 984.5 (976.0, 1004.5) | 996.1 (978.8, 1006.7)<br>n=10 | 993.8 (979.6, 1005.7)<br>n=24 | 0.581 |
| Native T1 >1050ms<sup>α</sup> | 0/8 (0) | 1/13 (7.7) | 0/29 (0) | 0.225 |
| Native T2 base (ms) | 47.3 (46.3, 48.0) | 47.8 (45.8, 48.3) n=10 | 47.6 (46.5, 49.2) n=27 | 0.776 |
| Native T2 mid (ms) | 47.6 (47.0, 48.0) | 48.0 (47.1, 49.5) n=9 | 47.7 (46.8, 48.8) n=25 | 0.676 |
| Native T2 >60ms<sup>α</sup> | 0/8 (0) | 0/10 (0) | 0/29 (0) | N/A |
| ECV Base (%) | 25.2 (24.3, 25.8) n=6 | 23.9 (20.8, 25.7) n=11 | 22.2 (20.9, 22.9) n=27 | 0.064 |
| ECV Mid (%) | 25.4 (22.1, 26.5) n=6 | 24.1 (23.3, 24.8) n=10 | 22.7 (21.5, 23.5) n=23 | 0.073 |
| ECV >30%<sup>α</sup> | 0/6 (0) | 1/13 (7.7) | 0/29 (0) | 0.396 |
| LGE Present | 3/7 (42.9) | 14/19 (73.6) | 1/29 (3.4) | **<0.001** |
| Total segments with LGE | 4/119 (3.4) | 40/323 (12.4) | 1/493 (0.2) | **<0.001** |

Data are presented as Median (Interquartile Range) or N (%). Patient group sizes as indicated in top row unless otherwise noted by denominator.
<sup>α</sup> Each count represents patient with value elevated above specified limit in either basal LV, mid LV, or region of interest

